# Predictors of Time-to-ART-Initiation Survival Times in a Random Sample of Adults Living with HIV from Malawi – A Historical, Nationally Representative Cohort Sample of 2004-2015 HIV Data

**DOI:** 10.1101/2024.01.04.24300777

**Authors:** Hemson Hendrix Salema

**Affiliations:** Warwick Medical School, University of Warwick

**Keywords:** HIV/AIDS, ART, ALHIV (Adults Living HIV), Survival analysis, Survival time, Cox proportional hazards model, ART-initiation, Universal test-and-treat

## Abstract

**Background:** Despite globally adapted universal test-and-treat (UTT) strategy of HIV management, survival time-to-antiretroviral-therapy initiation (TTAT) remains heterogenous and affected by diverse factors which remain unexplained in Malawi. This study explored correlates to TTAT-initiation in Malawi among adults living with HIV.

**Methods:** A multicentre retrospective cohort study was undertaken from eight centres. Medical records of (*n*=9,953) adult patients aged 15+ years old, were reviewed. A life table, the Kaplan-Meier log-rank, and Cox Proportion Hazard regression were used to calculate survival TTAT-initiation and its correlates, respectively. Adjusted Hazard ratio less than 1 (aHR <1) signified factors negatively associated, while aHR >1 meant factors positively associated with TTAT-initiation. Hazard ratio with 95% Confidence interval (95%CI) and *p*<0.05 were used to declare statistical significance.

**Results:** Data from (n=9,953) adult HIV patients were abstracted from hospital medical records. Patients median age was 40 (IQR: 33-48 years). 60.8% were females, 45.2% were younger adults of 20-39 years, and 78.8% were either married or cohabiting. 48.1% had advanced HIV disease; WHO clinical stage III, 24.5% had WHO stage IV, whereas 27.5% were asymptomatic; thus; 24.9% initiated ART due to low CD4+ count and 2.6% under PMTCT’s Option-B+. Findings from TTAT-initiation survival function analysis show that each patient had a single entry into the study and provided a total of 5,414 event-time-intervals, giving a 100% total event-failure without censored observations. Mean and median survival times were 527.2 days and 6 (IQR 0-5,414) days, respectively. Treatment-initiation (time at risk) was observed at the rate of 0.002 per 5,247,268 person-years. From multivariable Cox PH regression analysis, independent factors identified to be negatively associated with early (timely) antiretroviral treatment-initiation included; older age of 55+ years by 16% [aHR 0.84, 95%CI: (0.71–0.97)], male gender by 4% [aHR=0.96, 95%CI: (0.92–0.98)], bacterial causes by 5% [aHR=0.95, 95%CI: (0.89–0.99)], mycobacterial causes by 14% [aHR=0.86, 95%CI: (0.81–0.92)], high viraemia VL>1,000 copies/mL by 17% [aHR=0.83, 95%CI: (0.81–0.95)], registered in secondary tier and tertiary tier health facilities by 21% [aHR 0.79, 95%CI: (0.73-0.86) and 14% [aHR 0.86, 95%CI: (0.79-0.93) respectively, longer HIV survivorship (duration) by 58% to 85%, and having respiratory symptoms like coughing or breathlessness by 7% [0.93 95%CI: (0.88–0.97)]. These factors acted as barriers to early time-to-ART-initiation. In contrast younger adults of 20-39 years; [aHR=1.04, 95%CI: (1.02–1.19)], early mid-aged patients of 40-54 years; [aHR=1.03, 95%CI: (1.01– 1.21)], incomplete basic education level; [aHR 1.09, 95%CI: (1.02-1.20)], secondary education level; [aHR=1.06, 95%CI: (1.01-1.12)], Northern and Southern regions ethnicity; [aHR=1.07, 95%CI: (1.02–1.16)] and [aHR=1.06, 95%CI: (1.01–1.12) respectively, manifesting chronic headache or fevers; [aHR=1.12, 95%CI: (1.04–1.21)], being asymptomatic; [aHR=1.02, 95%CI: (1.02–1.13), (*p<*0.041)], being immunosuppressed with WHO clinical stage III; [aHR=1.86, 95%CI: (1.21-3.45)], WHO clinical stage IV; [aHR=2.80, 95%CI: (1.20-3.22)], protozoal pathological infection; [aHR=1.06, 95%CI: (1.02-1.15)], low CD4+ cell count <250 cells/µL; [aHR=1.05, 95%CI: (1.01–1.09), self-employment [aHR=1.04, 95%CI: (1.00–1.09)], and year of HIV diagnosis variable, were all positively associated with treatment-initiation and acted as precursors to early (timely) ART-initiation.

**Conclusion:** The study demonstrates that apart from meeting clinical eligible, different clinical and nonclinical factors contributed to time-to-treatment initiation among adults living with HIV. These factors; which are still prevalent in Malawi, have contributed to the spiralling and high mortality and morbidity from HIV/AIDS in Malawi and– hence, a knowledge of their existence, coupled with efforts to counteract and halt their occurrences, and strategies to strengthen and sustain the gained milestones in all tiers of health facility establishments across Malawi cannot be overemphasised.

## BACKGROUND

HIV remains the leading cause of mortality and morbidity worldwide and continues to disproportionately affect emerging economies; compromising population’s health of the world’s poorest people and swelling-up economic costs to their health systems. According to recent 2023 report of the Joint United Nations Programme on HIV/AIDS (UNAIDS) and the World Health organisation (WHO), by end of 2022, globally, 39 million people were living with HIV (PLHIV), 1.3 million became newly infected and 630, 000 people died from AIDS-related illnesses [1, 2]. Of the recent HIV epidemic, the WHO African region registered the greatest burden with 25.6 million PLHIV, 660,000 new infections, and 380,000 AIDS-related deaths [2]. Furthermore, an estimated 67% of the 38.4 million PLHIV worldwide in 2021 were from SSA [3]. The current HIV epidemic statistics bring the cumulative total to 85.6 million people ever infected and 40.4 million AIDS-related deaths since epidemic started [1]. Of all PLHIV in 2022, only 76% (29.8 million) were accessing antiretroviral therapy (ART) leaving 9.2 million people without access to ART.

Although still incurable, the untreatable virus; with its complex pathophysiology that demands complex treatment regimens [4], is no longer a “death sentence” as it has progressed from a fatal disease to a chronic illness owing primarily to the development of potent ART [5–7]. For over quarter a century, the combination antiretroviral therapy (cART) has been the global standard approach to treating HIV infection [8–10]. While the first cART regimens were inferior, the present ones are more potent, less toxic, and have fewer adverse-effects, have reduced pill burden – often one a day [11], or better still, a long-acting extended-release injectable ART regimen given every 1 or 2 months or every other month as 2 injections [12–18]. Moreover, they suppress HIV replication better, and have higher genetic barriers to resistance [11, 19]. Thus, present ART is highly effective in slowing HIV progression to AIDS, reduce AIDS-related mortality and morbidity [20, 21], and prevent further HIV transmission – as suppression of viral replication means achieving undetectable (viral) levels, and nontransmissible virus [22, 23], – however, ART-initiation time and coverage are principally crucial areas.

Timely ART initiation among PLHIV contributes a pivotal role to handle the epidemic. This then shifts the focus of care from survival to improving quality of life through adherence and retention in care and speed up of viral suppression [24] – and this defines an effective (HIV) care cascade. Thus, efficient HIV continuum of care demands timely (early) ART initiation for all PLHIV. Despite the fact, delayed or late ART start has been and remains a challenge among newly diagnosed HIV-seropositive and ART-naïve individuals. While immediate (early) ART-start following confirmed HIV diagnosis is now universal gold standard HIV management in the present universal test-and-treat (UTT) strategy [22, 25], previous management guidelines were to the contrary. Hitherto, potential PLHIV typically, were entered into care and observed moving down the HIV cascade of care continuum – a linear and unidirectional continuum of care [26–28] comprising of HIV testing, retention in pre-ART care, ART-initiation, and sustained suppression on ART – until they meet ART eligibility criteria by manifesting specific clinical features of advanced HIV disease (AHD) [29–34]. This strategy potentially promoted lost-to-follow-up and late ART-initiation.

Although ART-initiation is a crucial component of HIV cascade of care, barriers to timely treatment initiation and deficits in the spectrum of engagement in HIV care remains a realism. Earlier epidemiological evidence has demonstrated notable multiple factors as identifiable barriers that have a bearing on ART-initiation periods. Such distinct barriers include late HIV diagnosis, suboptimal linkage to, and retention in HIV care, insufficient ART usage and suboptimal ART adherence [35], infection route [36, 37], patients’ age [38–40], race, ethnicity, and environmental factors [41–43], gender [44], HIV subtypes, poor nutrition, co-infections, severe stress, one’s genetic background and immunological factors such as CD4 T-cell count [45–51], virological factors [52–57], psychosocial factors [58–63], and resource availability or socioeconomic factors [64]. These present significant impediments to achieving optimal treatment outcomes namely; undetectable viraemia and improved immune status [65]. Moreover, many PLHIV start and stop HIV care or ART multiple times over the course of their lives, creating the so-called “side door” re-entrance into the cascade for individuals who left the system [66] - this swells-up negative or undesirable treatment outcomes.

While the past decades have seen marked progressive decreases in HIV infection rates in developed nations, the developing world still lags behind. HIV continues to affect the world’s poorest populations and contribute to a cycle of poverty due to decreased productivity from acute to long-term comorbidities [67]. Within the low-middle-income countries (LMICs) the Sub-Saharan Africa (SSA) remains hugely disproportionally affected [1, 2].

Malawi, a relatively small, poor, and landlocked Sub-Saharan African nation in the Southern part of Africa, has one of the highest HIV prevalence globally; currently, estimated at 8.9% among adults [68]. With limited accessibility of care and services, SSA countries including Malawi, share the highest HIV/AIDS global burden [69]. Currently, Malawi HIV epidemic ranges from 4.0% to 14.2% [68]. In 2004 Malawi introduced free antiretroviral treatment to clinically eligible persons, however, not every eligible patient was able to promptly commence treatment potentially, resulting in consistently high AIDS-related deaths. In spite of high mortality attributed to delayed ART-initiation (even in the wake of free ART), factors contributing to the delays remain unexplained. This study aims to examine predictors associated with time-to-ART-initiation survival times since confirmatory HIV-seropositivity test results. While as presently there’re clear descriptions and cut-points to define early-, rapid-, same-day-, late-, and delayed ART-initiation – based on time of, vs condition at presentation to care and lag-time to starting treatment [70–73] previous guidelines and recommendations were based only on ‘standard of care’ where AIDS-related clinical events, and then, low CD4+ cell count were the reference points [71, 74–77]. All patients in the present (ADROC) study commenced ART following this strategy and this study explored correlates to treatment-initiation times in this cohort. Although presently 97.9% of PLHIV in Malawi are on cART [1, 2, 69] with as high as 88% same-day ART-initiation uptake [78], these achievements and milestones require to be improved further, strengthened, and be sustained and an exploration of correlates for or against time-to-ART-initiation can greatly contribute to this goal.

## METHODS AND MATERIALS

### Study Design, Setting, and Participants

The methodology employed in this study has already been [detailed somewhere in the ADROC Study](https://www.medrxiv.org/content/10.1101/2023.11.14.23298518v1). Briefly, a historical retrospective chart review cohort study was conducted between March and May 2016 in eight Health facilities randomly selected through multistage probability sampling across Malawi. Patients’ medical records (PMR) popularly called ‘ART Master cards’ – electronic medical records (EMR) or physical patients medical records (PPMR), were the sampling frames. HIV patient’s register and clinical progress notes were duly consulted for missing or additional information as necessary. Data collection sites included all existing tiers of health facility establishments in Malawi: thus, primary, secondary, and tertiary tier Health facilities – of which; 3 of the 8 research sites had PPMR. Follow-up time for each patient was calculated from HIV-seropositive confirmatory date to ART-initiation date. Of the eight data collection sites; one was in Northern Malawi, three in Central Malawi, and four in Southern region of Malawi.

### Study population: Inclusion & Exclusion Criteria

Enrolled participants were PLHIV aged 15+ years who were once commenced on ART following Malawi’s standardised treatment protocols between Jan. 2004 and Dec. 2015 – regardless of ART exposure duration, ART adherence, and patient’s status (alive or dead). All ART-naïve patients, and patients aged <15 years old were excluded from the study.

### Sample Size and Sampling Procedure

The study’s sample size and sampling procedure has been previously [detailed in another paper (the ADRCO Study)](https://www.medrxiv.org/content/10.1101/2023.11.14.23298518v1). The sample size was determined using double population proportion procedure used in previous studies and recommended in prevalence studies [79–84] using the following equation:

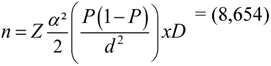

> **Where:** n = is the sample size,
>
> P = Prevalence of opportunistic disease in PLHIV in Malawi @10.6% HIV rate based on 2010 MDHS d = margin of error between sample and population sizes (n vs N) (0.006)
>
> 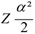 = statistical critical value at 95% confidence interval (1.96)
>
> D = design effect for a cluster sampling of sampling frames (DHOs, Health facilities & ART Master cards) estimated @ 2.0.

An allowance of 15% was applied to the estimated sample size (n=8,654) to compensate for poor documentation resulting in final sample size of (n=9,953). A three-stage probability sampling method was used to randomise the primary sampling units; the Health facilities and medical records data frames. Health facilities were selected from the list of 716 existing ART facilities as recorded in the 2015 Ministry of Health’s HIV Unit report [85, 86]. Participating sites were sampled using probability proportional to size from primary sampling units from the simple equation …. *K=N/n:*

> *…*where *…K* = is the sampling interval unit, …*N* = is the total cumulative population size (or the sampling frame (population), …*n* = is the number of sampling units to be selected [87–90].

At each research (ART) centre, exclusion criteria were applied to the primary data frame [ (*N*_1_) ], to form secondary sampling frame, [ *N* _2_ ]; a stratum of eligible patients. The sampling procedure was [then] replicated to randomly sample potential participants from *N* _2_. Anonymised or de-identified patients’ serial numbers [from (N _2_)] were listed in excel as a final sampling frame from which *n*_1_, *n*_2_, *n*_3_, *n*_4_, *n*_5_,………*n_x_* serial numbers were randomly selected. In research sites where patients’ records storage systems were electronic platforms and fully functional from the constructed sampling frame of eligible subjects ( *N* _2_ ), participants were randomly selected using computer-generated random sample in excel. The allocation of total study subjects at each health facility was based on the proportion of total number of ART-exposed adults.

### Data collection tools and procedures

Likewise, data collection procedure has been explained [somewhere in another study](https://www.medrxiv.org/content/10.1101/2023.11.14.23298518v1). Briefly, data were collected using a structured electronic tool purposefully designed and developed in Microsoft Access to capture data on both prevalent and incident AIDS-associated clinical conditions, associated risk factors, HIV history, ART history, and de-identified demographic details according to the ADROC Study variables. Prior to this, a pilot study was conducted from two ART sites to judge the useability, functionality, and validity and this gave chance to update or amend and realign the tools.

### Data Quality Assurance

The first exercise at each site was to elect and train a resident research team. The teams were strictly following the planned and approved data collection process. Training on data collection procedures, close supervision of research teams during data collection and random double data re-entry checks were implemented as part of quality control measures.

### Statistical analysis procedure

Statistical analyses were executed using Stata for windows version 15.1. Data were wrangled, cleaned, edited, categorised, and re-categorized prior to the actual analysis. Survival analysis involved (nonparametric) estimations of TTAT-initiation survival function with Kaplan-Meier (K-M) methods. Frequency and proportions were expressed as absolute numbers and percentages. A day was used as time scale to calculate time-to-ART-initiation. Person-days of observation was calculated as “date of ART initiation (event-function date) subtract from date of positive HIV diagnosis confirmation.” Descriptive survival analysis such as life table and K-M function curves were computed to estimate survival probability. The K-M Life Table was used to check probabilities of ART-initiation at different time intervals and to check cumulative survival probabilities. Survival function equality tests across different levels of variables (covariates) were tested using the log-rank test and survival (or hazard) function curves. Both bivariable and multivariable Cox Proportional Hazard (PH) regression model analysis was performed to investigate potential predictors of ART-initiation. For univariate (and bivariable) model, variables were purposefully selected based on their clinical significance. For multivariable Cox PH model, the predictors which revealed association with outcome variables at the univariable *p-*value (*p<*0.25) and the non-violation of proportionality test (*p>*0.05) determined variable inclusion into the final multivariable Cox PH regression model. Thus, the K-M curves were used to explore shapes of categorical predictors and determine their proportionality and inclusion in final multivariable model. The predictors’ fitness for the model was numerically verified using the Log-rank tests for equality across strata… [*model…1*]. In addition, the final model was double fitted using stepwise backward elimination of variables to judge the fitness of the model… [*model…2*]. Overall, results from [*model…1*] and [*mode…2*] were similar in both the number of variables and the effect size (ES) estimates. The final Cox PH model was then compared with the best of the parametric distribution models; the Weibull, Exponential, Gompertz, lognormal, through Cox-Snell residual reliability test judge the goodness-of-fit of model best-fitting the data. The Akaike’s Information Criterion (AIC) was used to evaluate the strength and validity of the parametric models that showed the smaller the value of the statistic; lowest AIC value, as best the fitting model. The Schoenfeld and scaled Schoenfeld residuals test was employed to assess final hazard regression model adequacy and numeric results were confirmed through scaled Schoenfeld residual curves. Hazard Ratios with 95% confidence intervals (HR, 95%CI) was computed and statistical significance was declared at 5% level (*p*<0.05). Cox proportional hazards regression model was used to determine factors associated with risk of time-to-ART-initiation by controlling confounding factors. Finally, the Cox-Snell residual plot was used to evaluate the overall goodness-of-fit of final Cox PH regression model.

### Operational definitions

**Survival time:** Length of time between HIV-seropositive date and ART-initiation date.

**Event-failure**: Antiretroviral therapy (ART)-initiation.

**Event date or Event-failure date:** Antiretroviral therapy (ART)-initiation date.

**HIV duration (survival):** Time from HIV diagnosis.

**Good adherence:** ≥95% adherence (missing only 1 out of 30 doses or 2 from the 60 doses).

**Fair adherence:** 85%–94% adherence (missing 2–4 out of 30 doses or 4–9 from 60 doses).

**Poor adherence:** <85% adherence (missing >5 out of 30 doses or >10 from 60 doses) [91].

**Educational level classification:** Based on UNESCO’s International Standard Classification of Education: incomplete basic education, basic education, secondary level, and tertiary level [92].

**Hazard ratio (HR): HR <** & **HR >:** Factors with HR <1 means negative association, HR >1 means positive association with time-to-ART-initiation.

## RESULTS

### Baseline socio-demographic characteristics

Sample disposition is presented in Table 2. Data for (*n*=9,953) enrolled participants were generated over an 11-year period – spanning from 2004 to 2015. The majority of participants were females (60.8%), young adults of 20-39 years (45.2%), and 78.8% were either married or cohabiting. Proportions of subjects with basic education and secondary education levels were indifferent; 33.7% & 32.8% respectively, and more than half (55.3%) were in paid employment. Educational level was categorized based on UNESCO’s International Standard Classification of Education: incomplete basic education, basic education, secondary level, and tertiary level [92]. 59.6% were recruited from Southern region of Malawi, 67.7% from tertiary (3^rd^ tier) Health facilities (i.e., Central hospitals ART clinics) and majority (65.1%, N=6,477) were urban residents.

All enrolled subjects were once commenced on ART under Malawi standardised treatment protocols. Most patients commenced treatment while in advanced HIV disease (AHD) phase; 48.1% and 24.5% had WHO clinical stage III & IV, respectively. 27.5% were asymptomatic at treatment initiation; of which 24.9% of patients had low CD4 count and 2.6% commenced ART under PMTCT’s Option-B+ criteria (were either breastfeeding or pregnant mothers).

CD4+ count, and viral load tests data were extracted on two time-points: as initial baseline and as follow-up test results – the latter were the most recent available test results. Overall, the mean (±SD) baseline initial CD4 count was 145.1±95.1 cells/µL. Most patients; 69.9%, had severe immunosuppression with baseline initial CD4 count <250 cells/µL. Similarly, the majority (96.4%), had severe viral failure with baseline initial viral load (VL) >1,000 copies/mL. The mean (±SD) baseline initial VL was 6.0±4.3 Log_10_ copies/mL, and majority had >4.9 Log_10_ copies/mL. 86.5% had lived with HIV for over 2 years, 78.4% had been on ART more than 2 years and 67.9% were continuing treatment with good adherence rate. Efavirenz-based Reverse Transcriptase Inhibitors (RTIs) ART-regimes were frequently used first-line treatment regimens in 67.1% of patients compared to 28.5% NVP-based regimens. On maintenance regimes; 95.4% were continuing with first-line regimens while 3.3% had been switched to second-line regimens; the boosted-Ritonavir PIs regimens – implying that they had proven first-line treatment failure.

### Survival Times-to-ART-Initiation

Descriptive findings of time-to-ART-initiation (TTAT) survival function show that each patient had a single entry into the study – giving a 100% total event-failure without censored observations. The study data were generated through a span of 11-year period and provided a total of 5,414 event-time-intervals, i.e., survival (failure) times. The mean and median survival times were 527.2 days and 6 (IQR 0-5,414) days, respectively. Treatment initiation; time at risk, was observed at the rate of 0.002 per 5,247,268 person-years. Time-to-ART-initiation incidence rate ratio by gender was 0.9477 (*p=0*.004).

The Kaplan-Meier (K-M) survival function curve for TTAT (Fig. 1A) demonstrates that survivorship function estimates descended steadily and swiftly the first 1,344 days (48 months) at which time, the survival probability was roughly 22%. It then tails-off sharply, and gradually reaching a 0% minimum value at 5,414 time-interval. Inversely, the Nelson-Aalen (N-A) cumulative hazard estimate curve (Fig. 1B) goes up, reaching maximum value at 5,414 time-intervals. Thus, the hazard function plot sharply curves (bends) up during the first 48 months (1,344 days) before gradually tailing off. The initial steep descent, or ascent, suggests that most patients started treatment shortly after HIV diagnosis. With the median survival of 6 days, this may further imply that for most patients, HIV diagnosis was made either while in advanced stages of the disease (a more probable reason), or that clinicians were proactively monitoring (following-up) treatment-naïve patients and start them on ART as soon as clinical eligibility was established.

**Figure 1 [A]:**
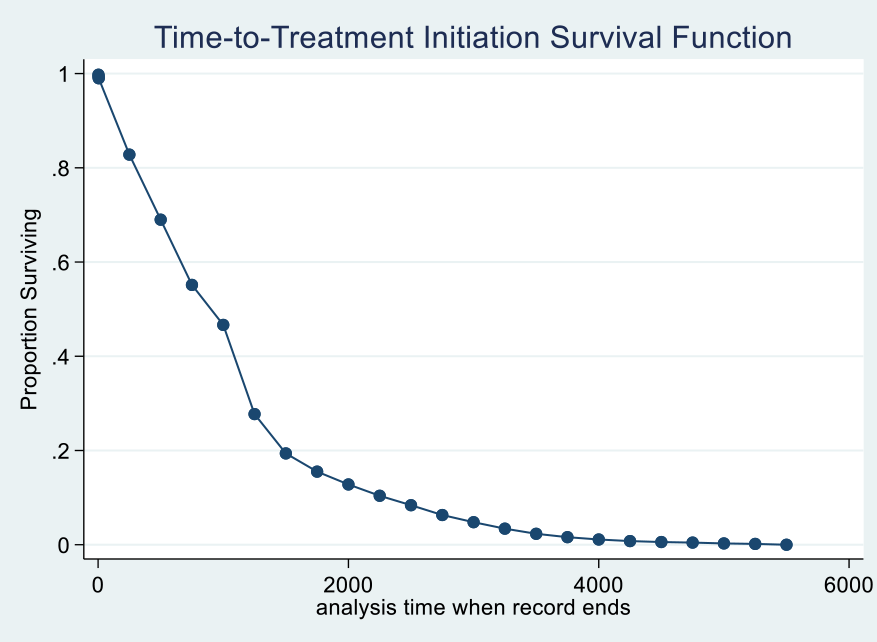
The K-M TTAT survival function. The Kaplan-Meier (K-M) survival function curve for time-to-ART-initiation demonstrates steady swift descent of the survivorship function estimates the first 1,344 days at roughly 22% survival probability before it tails-off sharply and gradually to 0% minimum value at 5,414 time-interval.

**Figure 1 [B].**
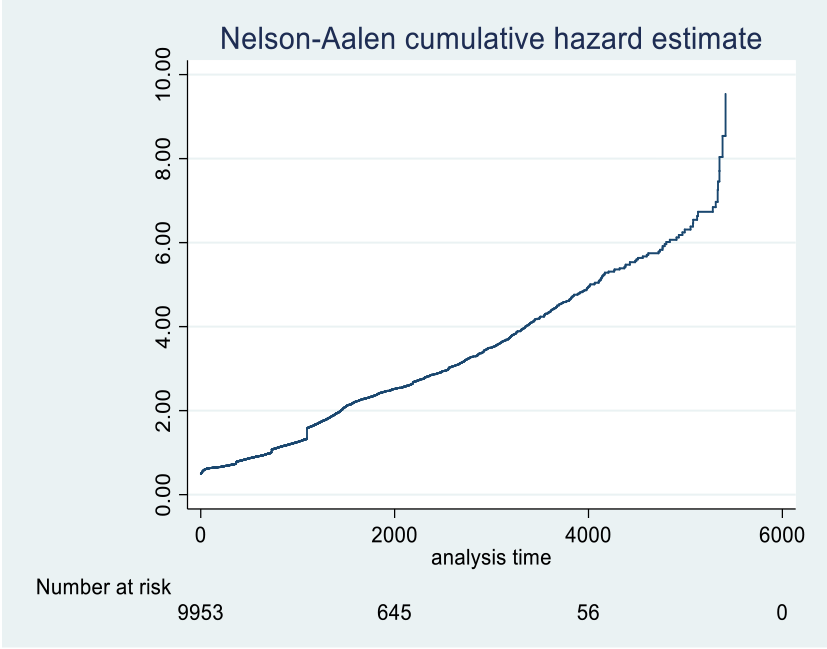
The Nelson-Aalen cumulative hazard estimate. The Nelson-Aalen cumulative hazard estimate does the opposite of the K-M curve, i.e., it curves up during the first 1,344 days (48 months) before gradually tailing off, reaching maximum value at 5,414 time-intervals.

The K-M Actuarial Lifetable (Table 1) demonstrates that the 6 days median survival time fell within the first time-interval and that almost half of all patients (N=4,912) were eligible for, and commenced ART by the end of 1^st^ time-interval. This implies that in these patients, treatment-initiation date was the same as HIV-diagnosis date (i.e., treatment was started on the same day or as rapidly as HIV diagnosis was made). Furthermore, based on the K-M actuarial lifetable results alongside (Fig. 3); over half of ART-initiation events (58.1%, N=5,779) occurred within the first 0-250 time-interval band (within the first eight-months). Fewer patients lived longer before being eligible for treatment such that, by 2,000^th^ and 4,000^th^ time-intervals - (i.e., 66.7 months or 5.6 years and 133.3 months or 11.11 years); only (N=645) and (N=56) patients respectively, were still [asymptomatic] and yet to be eligible for ART.

**Table 1:** The Kaplan-Meier Actuarial Life-Table illustrating proportions (of patients) surviving followed over treatment-initiation period. **Note:** Time-intervals are grouped into 250-interval-bands (in days), except for the first time-interval row. Over half of ART-initiation events (N=5,779) occurred within the first 0-250 time-interval band [meaning that 58.1% of patients’ clinical latency period fell within the first eight-months, (i.e., they became clinically eligible for ART – and were therefore commenced on ART within 8 months of HIV diagnosis]. Fewer patients lived longer before meeting ART eligibility criteria such that, by 2000^th^ and 4000^th^ time-intervals; [i.e., 66.7 months (5.6 years) and 133.3 months (11.11 years) post HIV diagnosis], only (N=645) and (N=56) patients respectively [Fig. 2], were still not eligible, hence, yet to commence treatment.

| Interval<br>(Time interval,<br>days) | Beginning Total<br>(No. at risk) | Failure (event)<br>(No. of events) | Censored<br>(lost)<br>observations | Survival<br>Function | Std. Error of the<br>estimated survival | [95% CI] |  |
| --- | --- | --- | --- | --- | --- | --- | --- |
| 1 2 | 9953 | 4912 | 0 | 0.5065 | 0.0050 | 0.4966 | 0.5163 |
| 0 250 | 9953 | 5779 | 0 | 0.4194 | 0.0049 | 0.4097 | 0.4290 |
| 250 500 | 4174 | 696 | 0 | 0.3494 | 0.0048 | 0.3401 | 0.3588 |
| 500 750 | 3478 | 698 | 0 | 0.2793 | 0.0045 | 0.2705 | 0.2882 |
| 750 1000 | 2780 | 428 | 0 | 0.2363 | 0.0043 | 0.2280 | 0.2447 |
| 1000 1250 | 2352 | 954 | 0 | **0.1405** | 0.0035 | 0.1337 | 0.1474 |
| 1250 1500 | 1398 | 421 | 0 | 0.0982 | 0.0030 | 0.0924 | 0.1041 |
| 1500 1750 | 977 | 195 | 0 | 0.0786 | 0.0027 | 0.0734 | 0.0840 |
| 1750 2000 | 782 | 137 | 0 | 0.0648 | 0.0025 | 0.0601 | 0.0698 |
| 2000 2250 | 645 | 121 | 0 | **0.0526** | 0.0022 | 0.0484 | 0.0572 |
| 2250 2500 | 524 | 101 | 0 | 0.0425 | 0.0020 | 0.0387 | 0.0466 |
| 2500 2750 | 423 | 105 | 0 | 0.0320 | 0.0018 | 0.0286 | 0.0355 |
| 2750 3000 | 318 | 77 | 0 | 0.0242 | 0.0015 | 0.0213 | 0.0274 |
| 3000 3250 | 241 | 69 | 0 | **0.0173** | 0.0013 | 0.0149 | 0.0200 |
| 3250 3500 | 172 | 55 | 0 | 0.0118 | 0.0011 | 0.0098 | 0.0140 |
| 3500 3750 | 117 | 36 | 0 | 0.0081 | 0.0009 | 0.0065 | 0.0101 |
| 3750 4000 | 81 | 25 | 0 | 0.0056 | 0.0007 | 0.0043 | 0.0073 |
| 4000 4250 | 56 | 17 | 0 | 0.0039 | 0.0006 | 0.0028 | 0.0053 |
| 4250 4500 | 39 | 10 | 0 | 0.0029 | 0.0005 | 0.0020 | 0.0041 |
| 4500 4750 | 29 | 6 | 0 | 0.0023 | 0.0005 | 0.0015 | 0.0034 |
| 4750 5000 | 23 | 9 | 0 | **0.0014** | 0.0004 | 0.0008 | 0.0023 |
| 5000 5250 | 14 | 5 | 0 | 0.0009 | 0.0003 | 0.0005 | 0.0017 |
| 5250 5500 | 9 | 9 | 0 | 0.0000 | . | . | . |

**Figure 2:**
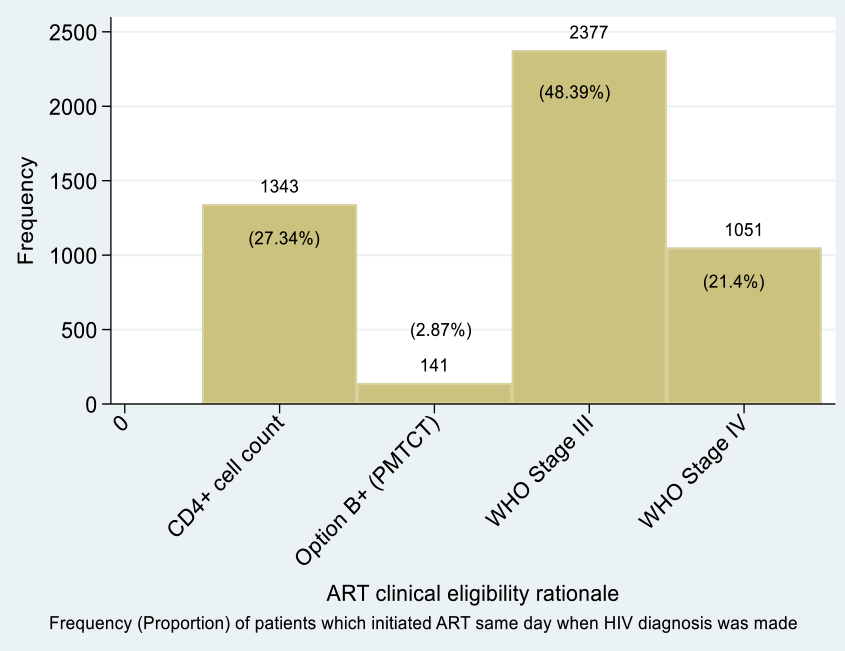
Distribution of Patients whose ART Dates were same as HIV Dates - (N=4,912): Graph of treatment eligibility criteria plotted against HIV dates and ART dates. Most patients with ART dates same as HIV diagnosis dates were symptomatic with AHD WHO clinical stages III and IV. Asymptomatic patients accounted for 30.2% with a considerable proportion (27.3%, N=1,343) commenced on ART under CD4 cell count criterion – hence, they were asymptomatic when treatment was commenced.

**Figure 3:**
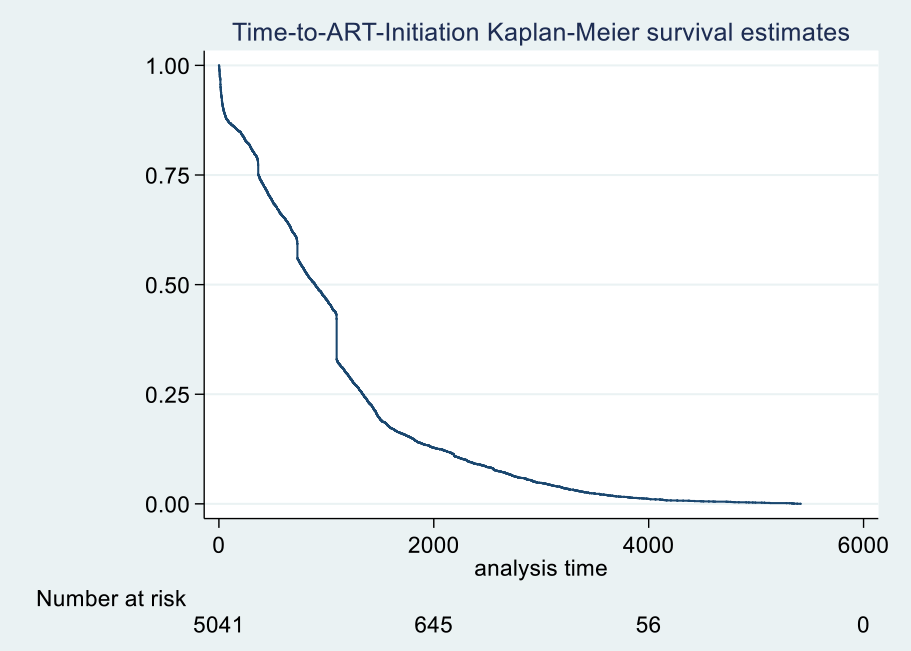
Overall Time-to-ART-Initiation K-M Survival Estimate (With At-Risk Table). The K-M curve for overall TTAT survival function with at-risk table (patients at-risk of failure) at specific time points, alongside Table 1, shows that the probability of surviving without ART decreased steadily. The survival rate of staying relatively healthy beyond 1,250th time-interval (i.e., 3.4 years), was only 14.1%. Survivorship probability between 6-10 years (deducible from Table 1) ranged from 5.3% down to 1.2%. At the final maximum time-interval [of 5000th time-interval mark (thus, 13.7 years)], only 0.14% subjects were likely to still be asymptomatic and stay relatively healthy without the need for antiretroviral-treatment.

### Predictors Associated with Time-to-ART-Initiation

In this study, time-to-ART-initiation (TTAT) survival function defines survival times elapsed between confirmatory HIV-seropositive diagnosis date and antiretroviral therapy eligibility/initiation date. Technically, TTAT-initiation is the [clinical latency period (asymptomatic or chronic HIV infection stage)](https://hivinfo.nih.gov/understanding-hiv/fact-sheets/stages-hiv-infection). For this survival analysis, the event-failure was a good and desirable occurrence, – thus, “risk of ART-initiation” precisely refers to time-to-event which is time-to-ART-initiation, *i.e.,* the probability of commencing ART. A positive risk means early (timely) ART-initiation while a negative risk means late or delayed ART-initiation without reference to number of days) lag-time). Factors with hazard ratio less than 1; (HR <1), means negative association with time-to-ART-initiation – i.e., these factors were associated with late (delayed) ART-initiation; hence, acted as barriers to early (rapid) or timely treatment initiation. Conversely, factors with HR >1 means positive association with time-to-ART-initiation; hence, being associated with early (rapid or timely) ART-initiation, henceforth, acted as precursors to rapid or timely ART-initiation.

To this end, in the multivariable Cox PH regression model adjusted for ART eligibility (Table 2), age was negatively associated with time-to-treatment initiation. The predictor demonstrated that every increase in age of 5 years, and 10 years, was negatively correlated with early ART-initiation by 2% and 4% respectively. Such that, compared to 15–19-year-olds, older age of 55+ years acted as a barrier to, and negatively associated with ART-initiation by 16% [aHR 0.84, 95%CI: (0.71–0.97)]. Male gender was 4% (aHR=0.96) negatively associated with early ART-initiation compared to females. On aetiological (pathological) factors responsible for causing ODs, the findings show that bacterial causes (aHR=0.95) and mycobacterial causes (aHR=0.86) were respectively, potential barriers to, and negatively associated with early ART-initiation, as was high viraemia VL>1,000 copies/mL (aHR=0.83). On health facility types as a factor influencing ART-initiation, secondary and tertiary healthcare facilities were associated with 21% [aHR 0.79, 95%CI: (0.73-0.86) and 14% [aHR 0.86, 95%CI: (0.79-0.93), negative association with early ART-initiation compared to primary health facilities. The findings also demonstrated that contraceptive use likewise acted as a barrier to rapid ART-initiation and was associated with delayed initiation by 7% - a different finding from South Africa that demonstrated no association with TTAT [93]. On HIV duration variable, the findings demonstrated that increase of every 5 and 10 years of HIV survivorship explained a 24% and 42% negative association to rapid ART-initiation. Thus, compared to patients with 1-2 years of living with HIV, those with 3-5 years, 6-10 years and >10 years HIV duration were 31%, 58% and 85% associated with delayed ART-initiation.

**Table 2:** Sample Disposition and Univariable and Multivariable Cox Proportion Hazard Regression Analysis of Time-to-ART-Initiation Survival Time.

| Sample Disposition:<br>Adult Living with HIV<br>on cART, 2004-2015 |  | Survival Regression Analysis of Time-to-ART-Initiation Survival Times |  |  |
| --- | --- | --- | --- | --- |
| Covariate (Variable) | Patients Distribution<br>Total N (%) | Univariable Analysis<br>(Crude estimates)<br>HR (95% CI) | Multivariable Analysis<br>aHR (95% CI) |  |
|  |  |  | Cox PH Distribution | Weibull Distribution |
| Age (years) |  |  |  |  |
| Age (years) | 9,953 (100%) | 1.00 (1.00–1.00) | 1.00 (0.99–1.00) | 1.00 (0.99–1.00) |
| Age_5 [age increase of every 5 yrs.] | 9,953 (100%) | 0.98 (0.97 – 0.99) | 0.98 (0.95 – 0.99) | 0.99 (0.97 – 1.01) |
| Age_10 [age increase of every 10 yrs.] | 9,953 (100%) | 0.96 (0.94 – 0.98) | 0.96 (0.90 – 0.98) | 0.98 (0.94 – 1.03) |
| Age group |  |  |  |  |
| 15-19 (Adolescents) | 358 (3.60) | 1 (ref) | 1 (ref) | 1 (ref) |
| 20-39 years (Young Adults) | 4,501 (45.22) | 1.03 (1.02–1.14)** | 1.04 (1.02–1.19)** | 1.05 (0.92–1.19) |
| 40-54 years (Early Mid-Aged) | 3,785 (38.03) | 0.97 (0.87–1.08) | 1.03 (1.01– 1.21)** | 1.01 (0.86–1.19) |
| 55+ years (Late Mid-Aged) | 1,306 (13.12) | 0.89 (0.79–0.92) | 0.84 (0.71–0.97)** | 0.99 (0.79–1.23) |
| Care type (Health facility type) |  |  |  |  |
| Primary care | 735 (7.38) | 1 (ref) |  |  |
| Secondary care | 2,482 (24.94) | 0.77 (0.71–0.83) | 0.79 (0.73–0.86)** | 0.69 (0.63–0.75) ** |
| Tertiary care | 6,736 (67.68) | 0.85 (0.78–0.91) | 0.86 (0.79–0.93) ** | 0.81 (0.74–0.87) ** |
| Residential areas |  |  |  |  |
| Rural | 2,047 (20.57) | 1 (ref) |  |  |
| Semi-urban | 1,427 (14.34) | 1.10 (1.03–1.17) | 1.08 (1.00–1.15) ** | 1.11 (1.03–1.19) ** |
| Urban | 6,477 (65.08) | 1.07 (1.02–1.13) | 1.06 (1.01–1.12) ** | 1.10 (1.05–1.16) ** |
| Regional areas (geographical region) |  |  |  |  |
| Central Region | 2,441 (24.53) | 1 (ref) |  |  |
| Northern Region | 1,585 (15.92) | 1.08 (1.02–1.15) | 1.07 (1.02–1.16) ** | 1.20 (1.13–1.29) ** |
| Southern Region | 5,927 (59.55) | 1.01 (1.01–1.06) | 1.06 (1.01–1.12) ** | 1.05 (1.01–1.11) ** |
| Gender |  |  |  |  |
| Female | 6,055 (60.84) | 1 (ref) | 1 (ref) | 1 (ref) |
| Male | 3,898 (39.16) | 0.96 (0.93–0.98) | 0.96 (0.92–0.98)** | 0.97 (0.92–1.02) |
| Marital status |  |  |  |  |
| Previously married | 1,380 (13.87) | 1 (ref) | 1 (ref) | 1 (ref) |
| Married (partnership) | 7,837 (78.74) | 1.02 (1.01–1.08) | 1.00 (0.94–1.06) | 1.02 (0.96–1.08) |
| Single (never married) | 733 (7.36) | 1.02 (1.01–1.15) | 0.98 (0.88–1.08) | 0.97 (0.88–1.07) |
| Education level |  |  |  |  |
| Tertiary education | 713 (7.16) | 1 (ref) | 1 (ref) | 1 (ref) |
| Incomplete basic education | 3,349 (33.65) | 1.14 (1.05–1.24) | 1.09 (1.02–1.20)** | 1.09 (1.00–1.20) |
| Basic education. | 3,260 (32.75) | 1.04 (1.02–1.10) | 1.04 (0.98–1.10) | 1.03 (0.97–1.09) |
| Secondary education. | 2,631 (26.43) | 1.07 (1.02–1.13) | 1.06 (1.01–1.12)** | 1.05 (0.99–1.11) |
| Employment status |  |  |  |  |
| Salaried-employees | 5,506 (55.32) | 1 (ref) | 1 (ref) | 1 (ref) |
| Self-employed | 4,447 (44.68) | 1.02 (1.00–1.06) | 1.04 (1.00–1.09)** | 1.03 (0.99–1.08) |
| Occupation type |  |  |  |  |
| Manual occupations | 1,658 (16.66) | 1 (ref) | 1 (ref) | 1 (ref) |
| Other (menial) occupations | 4,949 (49.72) | 0.98 (0.93–1.04) | 0.99 (0.93–1.05) | 1.00 (0.95–1.07) |
| White collar | 3,346 (33.62) | 0.96 (0.90–1.02) | 0.99 (0.92–1.05) | 0.99 (0.93–1.06) |
| ART clinical criteria |  |  |  |  |
| CD4 cell count | 2,475 (24.87) | 1 (ref) |  |  |
| Option-B+ | 261 (2.62) | 1.01 (0.88–1.14) | 0.94 (0.79–1.12) | 0.89 (0.78–1.02) |
| WHO Stage III | 4,781 (48.04) | 0.95 (0.91–1.03) | 1.86 (1.21–3.45)** | 0.50 (0.13–2.04) |
| WHO Stage IV | 2,436 (24.48) | 0.92 (0.87–0.97) | 2.80 (1.20–3.22)** | 0.43 (0.11–1.72) |
| Aetiological agent |  |  |  |  |
| Asymptomatic/universal | 2,732 (27.45) | 1 (ref) | 1 (ref) | 1 (ref) |
| Bacterial conditions | 1461 (14.68) | 0.91 (0.86–0.97) | 0.95 (0.89–0.99)** | 0.90 (0.84–0.96) |
| Fungal (mycotic) cond. | 711 (7.14) | 0.94 (0.86–1.03) | 0.97 (0.89–1.06) | 0.94 (0.86–1.02) |
| Lymphomatous conditions | 216 (2.17) | 0.96 (0.84–1.10) | 0.99 (0.86–1.15) | 0.98 (0.85–1.13) |
| Mycobacterial cond. | 3,529 (35.46) | 0.94 (0.89–0.98) | 0.86 (0.81–0.92)** | 0.92 (0.87–0.97) |
| Parasitic (protozoan) | 366 (3.68) | 1.01 (0.90–1.13) | 1.06 (1.02–1.15)** | 1.03 (1.01–1.16) |
| Viral conditions | 938 (9.42) | 0.98 (0.92–1.06) | 1.03 (0.95–1.11) | 0.99 (0.91–1.06) |
| CD4 count initial (recent) |  |  |  |  |
| CD4 count( _initial) | 9,953 (100%) | 0.99 (0.99–1.00) | 1.00 (1.00–1.00) | 0.99 (0.98–0.99)** |
| CD4_100i | 9,953 (100%) | 0.98 (0.96–1.00) | 1.00 (0.98 – 1.04) | 1.00 (0.98 – 1.04) |
| CD4_250i | 9,953 (100%) | 0.95 (0.91–1.01) | 1.02 (0.96 – 1.09) | 1.02 (0.96 – 1.09) |
| CD4 count initial |  |  |  |  |
| >250 cells/μL | 6,956 (69.89) | 1 (ref) | 1 (ref) | 1 (ref) |
| <250 cells/μL | 2,988 (30.02) | 1.05 (1.01–1.10)** | 1.15 (1.10–1.19)** | 1.15 (1.10–1.19)** |
| Covariate (Variable) | Patients Distribution<br>Total N (%) | Univariable Analysis<br>(Crude estimates)<br>HR (95% CI) | Multivariable Analysis<br>aHR (95% CI) |  |
|  |  |  | Cox PH Distribution | Weibull Distribution |
| Initial VL levels |  |  |  |  |
| VL initial | 9,953 (100%) | 1.00 (1.00–1.00) | 1.00 (0.99–1.00) | 1.00 (1.00–1.00) |
| VL_1000i (1,000) | 9,953 (100%) | 0.99 (0.99–0.99) | 1.00 (0.99–1.00) | 1.00 (0.99–1.00) |
| VL_10,000i (4.0 Log<sub>10</sub>) | 9,953 (100%) | 0.99 (0.99–0.99) | 1.00 (0.99–1.00) | 1.00 (0.99–1.00) |
| VL_100,000i (5.0 Log<sub>10</sub>) | 9,953 (100%) | 0.99 (0.99–0.99) | 1.00 (0.99–1.00) | 1.00 (0.99–1.00) |
| VL_1,000,000i (6.0 Log<sub>10</sub>) | 9,953 (100%) | 0.95 (0.93–0.97) | 1.01 (0.98–1.03) | 1.02 (0.97–1.03) |
| Initial VL levels categorisation |  |  |  |  |
| <1,000 copies/mL | 355 (3.57) | 1 (ref) | 1 (ref) | 1 (ref) |
| >1,000 copies/mL | 9,590 (96.35) | 0.89 (0.80–0.99) | 0.83 (0.81–0.95)** | 0.95 (0.85–0.96)** |
| Contraception |  |  |  |  |
| Yes | 7,087 (71.20) | 1 (ref) | 1 (ref) | 1 (ref) |
| No | 2,866 (28.80) | 0.95 (0.92–1.07) | 0.93 (0.88–0.98)** | 0.92 (0.88–0.97)** |
| HIV Diagnosis year |  |  |  |  |
| Jan 1990-Dec 2003 | 1636 (16.44) | 1 (ref) | 1 (ref) | 1 (ref) |
| Jan-Dec 2004 | 439 (4.41) | 1.24 (1.12–1.39)** | 1.25 (1.12–1.39)** | 1.27 (1.14–1.42)** |
| Jan-Dec 2005 | 1122 (11.27) | 1.28 (1.18–1.39)** | 1.29 (1.20–1.40)** | 1.28 (1.19–1.39)** |
| Jan-Dec 2006 | 1270 (12.76) | 1.27 (1.18–1.37)** | 1.27 (1.18–1.37)** | 1.28 (1.19–1.38)** |
| Jan-Dec 2007 | 1250 (12.56) | 1.21 (1.13–1.31)** | 1.22 (1.14–1.32)** | 1.24 (1.15–1.33)** |
| Jan-Dec 2008 | 819 (8.23) | 1.17 (1.07–1.29)** | 1.23 (1.12–1.35)** | 1.24 (1.13–1.36)** |
| Jan-Dec 2009 | 482 (4.84) | 1.26 (1.15–1.38)** | 1.33 (1.21–1.45)** | 1.36 (1.24–1.48)** |
| Jan-Dec 2010 | 490 (4.92) | 1.25 (1.14–1.37)** | 1.32 (1.20–1.45)** | 1.29 (1.17–1.41)** |
| Jan-Dec 2011 | 709 (7.12) | 1.26 (1.16–1.38)** | 1.36 (1.24–1.48)** | 1.39 (1.28–1.52)** |
| Jan-Dec 2012 | 648 (6.51) | 1.38 (1.26–1.52)** | 1.88 (1.65–2.14)** | 2.02 (1.78–2.30)** |
| Jan-Dec 2013 | 581 (5.84) | 1.49 (1.30–1.61)** | 1.94 (1.69–2.24)** | 2.13 (1.85–2.46)** |
| Jan-Dec 2014 | 371 (3.73) | 1.61 (1.39–1.85)** | 2.15 (1.82–2.55)** | 2.35 (1.99–2.78)** |
| Jan-Dec 2015 | 136 (1.37) | 1.49 (1.19–1.88)** | 2.02 (1.58–2.59)** | 2.14 (1.67–2.74)** |
| ART period |  |  |  |  |
| Early-ART period | 9,953 (100%) | 1 (ref) | 1 (ref) | 1 (ref) |
| late-ART period |  | 1.04 (0.98–1.08) | 0.73 (0.67–0.80) ** | 0.70 (0.64–0.76) |
| HIV Duration (days/years) |  |  |  |  |
| HIV duration (days) | 9,953 (100%) | 0.99 (0.99–0.99) | 0.99 (0.99–0.99)** | 0.99 (0.99–0.99)** |
| HIV duration_5years | 9,953 (100%) | 0.74 (0.70 – 0.77) | 0.76 (0.73 – 0.81)** | 0.73 (0.72 – 0.81)** |
| HIV duration_10years | 9,953 (100%) | 0.55 (0.50 – 0.59) | 0.58 (0.53 – 0.64)** | 0.54 (0.51 – 0.64)** |
| HIV Duration groups (years) |  |  |  |  |
| 1-2 years | 1,342 (13.48) | 1 (ref) |  |  |
| 3-5 years | 2,610 (26.22) | 0.68 (0.63–0.72) | 0.69 (0.64–0.73) ** | 0.37 (0.35–0.40) |
| 6-10 years | 4,440 (44.61) | 0.41 (0.38–0.44) | 0.42 (0.39–0.44) ** | 0.22 (0.21–0.24) |
| >10 years | 1,561 (15.68) | 0.16 (0.14–0.17) | 0.15 (0.14–0.17) ** | 0.10 (0.10–0.11) |
| Clinical presentation to care |  |  |  |  |
| HTC/VCT, Option B+ | 4173 (41.93) | 1 (ref) |  |  |
| Cough/breathlessness | 2389 (24.0) | 0.96 (0.89–1.05) | 0.93 (0.88–0.97)** | 1.00 (0.95–1.06) |
| Diarrhoea or weight loss | 725 (7.28) | 0.93 (0.86–0.99) ** | 0.99 (0.90–1.10) | 0.96 (0.88–1.04) |
| Fever & headache | 1157 (11.62) | 1.08 (1.03–1.14) ** | 1.12 (1.04–1.21)** | 0.98 (0.92–1.05) |
| Other symptoms | 1079 (10.84) | 1.03 (0.96–1.11) | 1.02 (1.02–1.13)** | 0.99 (0.92–1.07) |
| Skin or oral lesions | 430 (4.32) | 1.01 (0.91–1.12) | 1.01 (0.87–1.17) | 1.03 (0.93–1.14) |

Conversely, the multivariable Cox PH predictive model demonstrated multiple factors positively associated with time-to treatment-initiation and acted as precursors to timely (early) ART-initiation. Compared to the younger population aged 15-19 years, patients aged 20-39 years, and 40-54 years old, were associated with rapid ART-initiation by 4% and 3%, respectively [aHR=1.04, 95%CI: (1.02-1.19)] and [aHR=1.03, 95%CI: (1.01-1.21)]. On patient’s education levels, compared to tertiary education incomplete basic education, and secondary education explained a 9% and 6% positive association with rapid ART-initiation [aHR 1.09, 95%CI: (1.02-1.20) versus aHR=1.06, 95%CI: (1.01-1.12)] respectively. This association was however not reported from a recent study from Africa which observed no correlation [93]. Comparably, the likelihood of rapid ART-initiation was increased by 7% in Northern region and by 6% in Southern region compared to the Central region of Malawi (*p=*0.001). On clinical symptoms at presentation of care, time-to-ART-initiation was increased by 12% in patients presenting with chronic headache and/or fevers compared to asymptomatic patients (*p<*0.041). Consequently, manifesting with AHD: WHO stage III was positively associated with early ART-initiation by 86% (aHR=1.86, 95%CI: (1.21-3.45) whereas WHO stage IV explained a 3-fold increase in early ART-initiation (aHR=2.80, 95%CI: (1.20-3.22). Similarly, protozoal causes (aHR=1.06, 95%CI: (1.02-1.15), low CD4 count <250 cells/µL [aHR=1.05, 95%CI: (1.01–1.09) and self-employment [aHR=1.04 (1.00–1.09) were all positively associated with rapid ART-initiation with varying ES. Significant positive associations were also observed in year of HIV diagnosis variable (from 2004 through to 2015). Overall, the analysis demonstrated that hazard rates were progressively increasing in stepwise pattern throughout the 11-year-period data were generated except in 2007 & 2008 (Fig. 4). Early (rapid) ART-initiation increased by 25% among patients diagnosed with HIV in 2004 and by 88% in 2012 (*p<*0.0001). Moreover, the rates were over 2-fold higher in 2014 and 2015; thus, [aHR 2.15, 95%CI: (1.82–2.55) and aHR 2.02, 95%CI: (1.58–2.59) respectively.

**Figure 4:**
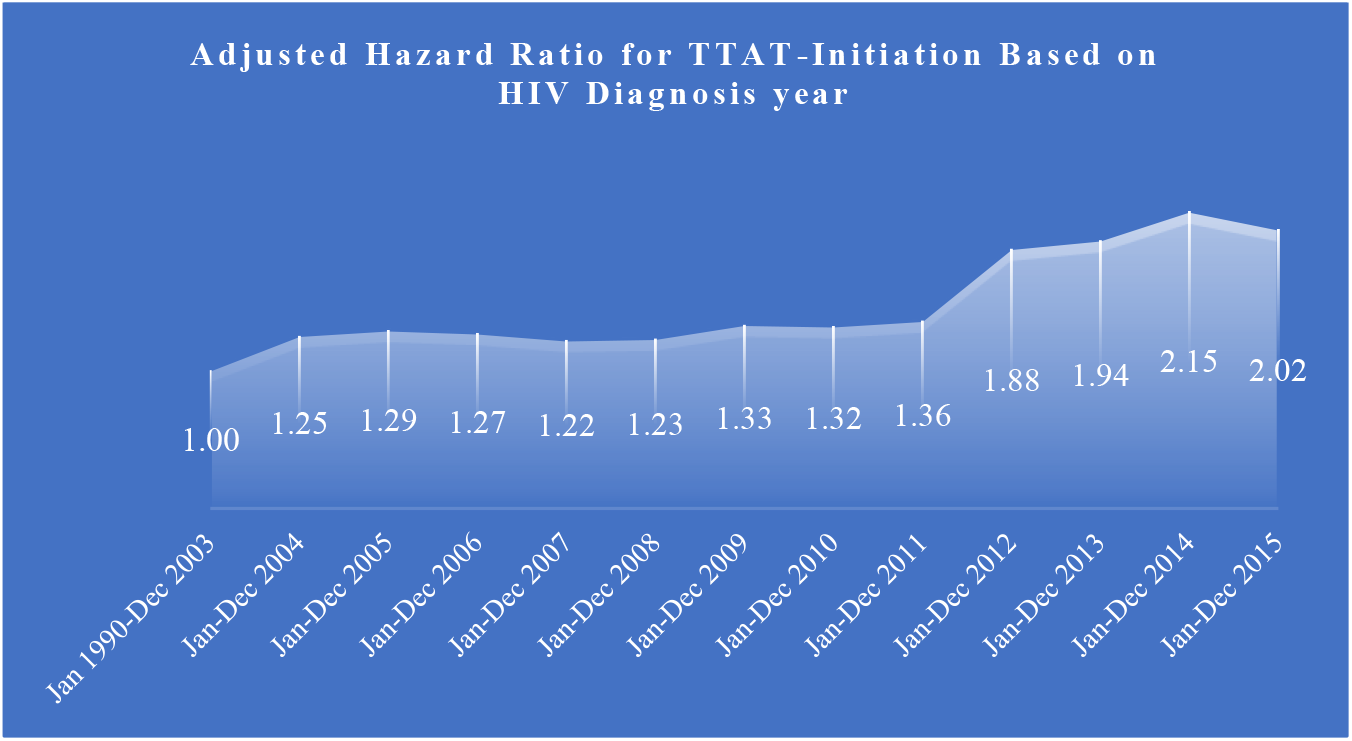
Adjusted Hazard Ratios for Time-to-ART-Initiation based on HIV Diagnosis year. Associations of HIV diagnosis year with TTAT (2004-2015). Overall, from 2004, adjusted hazard ratios, progressively increased in a stepwise pattern through to 2015 except in 2007-2008. The highest peak was in 2014.

## DISCUSSION

This study, which to my knowledge is the first of its kind in Malawi to analyse ART-initiation predictors among treatment-exposed adults using panel data, has demonstrated associations between demographical, socio-economical, geographical, behavioural, and clinical factors with time-to-ART-initiation. Patients were initiated on treatment at different periods between January 2004 and December 2015 and followed till death or otherwise lost-to-follow-up, stopped, or defaulted treatment. Globally, overall, there’s dearth of panel data studies similar to the present one – except a few with distinctive methodological differences that limit direct comparisons with the present one. Notwithstanding, comparisons are drawn from epidemiological studies with similar aims to the present one especially those that explored factors associated with late (delayed) ART-initiation in clinically eligible individuals. In this nationally representative sample, 100% treatment-initiation rate was observed at the rate of 0.002 per 5,247,268 person-years and time-to-ART-initiation incidence rate ratio by gender was 0.9477 (*p=*0.004). The median (IQR) survival time-to-ART-initiation in the present study was 6 (IQR: 0-5414) days. This finding is similar to 5 (IQR 1-13) days median time from West African countries – Burkina Faso, Cote d’ Ivoire, Mali, and Togo, [94], and similar to 4 (IQR 1-9) days from Ethiopia study [95] but was much lower than 29 (IQR: 0-514) days from UK study [73], lower than 21 (IQR: 17-39) days from Portugal [96] and from Republic of South Africa (RSA) [97] and, than 31 (QI-Q3: 8-537.5) days in Croatia [98]. Furthermore, in another RSA study, median time to ART-initiation was 12 (IQR: 1-59) days [99]. A multicentre European cohort study reported a median time from OD diagnosis to ART-initiation of 30 (IQR:16-58) days [100]. These findings highlight the fact that time-to-ART-start remain universally heterogenous and relatively longer even in settings with advanced and effective health service like Europe. In a multivariable Cox PH model adjusted for time-to-ART-initiation, structural factors related to demographic, clinical, and socio-economic marginalisation of PLHIVA, posed important influences on ART-initiation.

### Demographic Correlates to Time-to-ART-initiation

Demographic variables in the present study included age, gender, marital status, and family relation. The findings demonstrate that age, family (guardian) relation, and gender were associated with time-to-ART-initiation in a multivariable predictive model with different effect size (ES) estimates. Thus, late mid-age; 55+ years, was a barrier to, and negatively associated with early ART-initiation by a factor of 16%. Inversely, younger adults (20-39 years), and early mid-aged adults (40-54 years), were positively associated with early ART-initiation. This study’s findings on advancing age as barrier to early ART-initiation appear lower than findings from Canada study which reported a 40% positive association between old age [advancing age] and late (or delayed) ART-initiation [35], but are higher than 3% positive association reported more recently from another Canada study [71]. On the contrary, this finding is inverse relation to another study; also from Canada, which reported 2% positive association between aging and rapid-ART-initiation [101]. The current study finding on younger age Vs TTAT are in congruent with West African multinational study reported that younger age (of at least 24 years) was associated with early ART-initiation [94] – a similar association was also observed from South African KwaZulu Natal study [102]. The KwaZulu Natal study reported that early mid-age (of 35-44 years) had a 63% positive association with early ART-initiation (in males) and 90% probability (in females) whereas being ≥45 years old was a barrier to early ART-initiation [102] – the findings also shared in a study from Iran where old age was reported to be significantly associated with risk of late ART-initiation [aOR:1.02, 95%CI:(1.00-1.04)] [103] and a similar pattern was also observed from China [104].

The study also demonstrated that males were associated with delayed ART-initiation. This finding, however, differs with studies from Canada (Joseph, *et al.,* 2016) and the US [105] where no association between gender and time-to-ART-initiation was observed and from southern Iran study which reported no association between gender and treatment initiation times [103]. However, studies from Zimbabwe [106], Ethiopia [107], and Cameroon [108], reported that male gender was a risk factor for late-ART-presentation – therefore, a barrier to rapid treatment-initiation. Contrastingly, a study from Mozambique reported that female gender was associate with lower risk of late ART-initiation [109]. In addition, an RSA study exploring determinants of time-to-ART-initiation observed similar to the present study that hypothesised that females were associated with early ART-initiation – i.e., females had higher probability to rapid ART-initiation [(aHR=3.12-fold, 95%CI:2.87–3.38, *p*< 0.0001), (aOR=11.92-fold, 95%CI:( 9.81–14.52), *p*< 0.0001)] [99] likewise a study from China [104]. The gender difference observed in the present study and from studies from other SSA nations in comparison to those from developed nations like the US highlight the fact that within developing nations, males have less health-seeking behaviours than females [110, 111]. Across the globe, prevailing concepts of masculinity lead to men being less likely than their counterparts to seek healthcare, less likely to have HIV testing and less likely to initiate and adhere to ART [112]. This challenge is arguably more prevalent in SSA region where men and boys with HIV are 20% less likely to know their status and 27% less likely to access ART than their counterparts [113]. Although marital status was not statistically significantly associated with TTAT-initiation in the present study, the predictor has been established as an independent factor elsewhere [44] with reports that being married is associated with lower risk of late ART-initiation [109]. The Iran study exploring factors associated with late ART-initiation observed that being single was associated with late ART start by 80% (aOR: 1.8, 95%CI:1.17-2.78) [103] as was also reported in China [104]. This study also found that dependency on guardian escort was 9% negatively associated with early ART-initiation. This type of correlation is typical to this study as it has not been examined elsewhere. Nonetheless, this finding may be attributed to dependency behaviour widely common in most communities [114].

### Socioeconomic & Geographic Correlates to Time-to-ART-initiation

Socioeconomic variables in this study included education level, employment status, occupation classification, residential status, and geographical region. Different socioeconomic factors often interrelate to, and influence each other. Thus; one’s employment dictates their income whose level often correlates to their education attainment which often helps to dictate their employment. Moreover, geographic, and residential status often dictates one’s income level. These in turn influence the general public health status and health service delivery and operations. However, this study demonstrated mixed findings that patient’s education level and employment status were associated with early (timely) ART-initiation. Thus, having either incomplete basic education and, having secondary education, were positively associated with early ART-initiation by 9% and 6% respectively, as was self-employment (4%). These findings compare with the Mozambique study which reported that secondary or tertiary education had lower risk of late ART-initiation [109]. The finding however contrasts with studies from Canada [101], and Ethiopia [107] both of which found no statistically significant association between education status and rapid time-to-treatment-initiation. on the effect of employment status, a West African study, moreover, reported that people who “self-perceived” to be in (very) difficult financial situation were highly associated with early ART-initiation (aHR=1.44) [94]. According to socioeconomic-education level relationship, these people possess expectedly incomplete basis or basic education levels. However, conversely, participants with wealth index within Quantiles 2-5 were found to be statistically significantly associated with 38% higher probability of early ART-initiation in a logistic regression [93].

Geographical factors significantly associated with TTAT-initiation in this study included type of health facility and specific data collection centres. However, the associations, whether positive or negative, appear to depend on health facility tier and geographical location. Thus, secondary tier, and tertiary tier Health facilities overall, acted as barriers to, and were negatively associated with early ART-initiation by 21% and 14%. On the other hand, data collection centre variable significant findings suggest that Nkhoma ART centre (a semi-urban, secondary tier facility) was associated with early ART-initiation by 54% (aHR=1.54), and DREAM programme ART centre (an urban, primary ART centre) by 12% (aHR=1.12). Inversely, Mzuzu Central hospital (an urban ART centre), and Thyolo District hospital (a semi-urban ART centre), were 43% and 47% respectively, significantly associated with delayed ART-initiation. The geographical factors positive association findings in this study appear consistent with findings from Cameroon [108] and Canada [35] that explored factors associated with late ART-initiation. A Montreal (Canada) study exploring geographical factors associated with late ART-initiation reported that type of health facility; thus, university medical centres [a 3^rd^ tier university teaching hospital] versus community facilities [primary tier health facility], were independently associated with early ART-initiation [aOR=2.03] [115]. In addition, two more studies from Canada both demonstrated significant positive associations between patient’s residential status and rapid ART-initiation [71, 101]. In another setting, Ivana & colleagues (2022) in the ECHO Trial involving women from four African nations observed that one’s geographical location had an influence on TTAT. Thus, the ECHO Trial observed that compared to RSA, participants from Zambia were 2.73-fold (95%CI:1.21-6.11) highly likely to rapid ART initiate [93]. Similarly, geographical location was likewise observed to be highly associated with rapid ART-initiation; thus, compared to Lome (Togo), the hazard (aHR) for rapid ART-initiation was 5.15-fold for Abidjan (Côte d’Ivoire), 4.36-fold for Bamako (Mali), and 7.77-fold for Ouagadougou (Burkina Faso) [94]. Moreover, a multicentre study of two European nations; UK (London) and France (Paris) assessing factors associated with time between OD diagnosis and ART-initiation observed that patients in London were associated with longer time to ART-initiation by 38% (aHR=0.68, 95%CI:0.48-0.80) when compared to patients from Paris [100]. This finding demonstrates that geographic location has a bearing on treatment initiation time and compares with the finding in the present study.

On the other hand, on patients’ residential status, the present (ADROC) study found that compared to rural area, semi-urban and urban areas explained 8% [95%CI: 1.00-1.15], and 6% [95%CI: 1.01-1.12] respectively, positive association to early (timely) ART-initiation. This finding compares with but is much lower than 3.93-fold reported from Ethiopia where urban living was positively associated with early (same-day) ART-initiation [116] and is also lower than 2.02-fold [(95%CI: 1.37– 2.97), p<0.001] reported also from Ethiopia [95]. Notwithstanding, the study findings on residential location differs with one from Brazil which investigated the effect of socio-economic inequalities using education level as a proxy, on late initiation of ART-initiation [117]. This study demonstrated that compared to tertiary education, incomplete basic education was 89% [aOR:1.89, 95%CI:1.47–2.43)], basic education was 61% [aOR:1.61, 95%CI:1.23–2.10)], and secondary education was 35% [aOR:1.35, 95%CI:1.09–1.67)], increased odds of late ART-initiation [117].

Variations on geographical status findings in this study could be explained by variances in health service delivery, health facility’s patient-load, health facility geographical location, and population density between full (complete) and partial public sector health facilities. Thus, DREAM programme and Nkhoma ART medical centres are not-for-profit semi-public sector health facilities run by mission institutions under the Christian Health Association of Malawi (CHAM). These have at least adequate health personnel and relatively lower patient-load opposed to full public sector hospitals such as Mzuzu and Thyolo. Furthermore, historically, the Northern Region of Malawi is less populated while the South is denser than the Central. And, the North is less developed than Central and South Malawi – hence, do not attract many professionals, therefore, experience high disproportional patient-clinician ratio that affect the rate of health service delivery including HIV management. On the other hand, Thyolo District in Southern region has had one of the highest HIV rates historically [118, 119]. Finally, Southern Malawi has many health service delivery non-governmental organisations (NGOs). Often, these factors have attributed combined synergy for either improved or delayed ART-initiation.

### Bio-Clinical Correlates to Time-to-ART-initiation

Bio-Clinical predictors in this study included patient’s presenting features, aetiological agents responsible for ODs, patient’s CD4+ count and HIV-RNA load levels, ART eligibility criteria, ART regimen formulations, prophylaxis treatment for ODs, HIV duration, clinical latency period and HIV diagnosis year.

The current study demonstrated that chronic cough and/or breathlessness (7%), chronic diarrhoea and/or weight loss (10%), and chronic (or intermittent) fevers and headaches (14%), were negatively associated with rapid ART-initiation, and acted as barriers to treatment, hence, associated with delayed ART-initiation. Similarly, on aetiological agent variable, bacterial, and mycobacterial aetiological causes were negatively associated with rapid-ART-initiation by 5% and 14%, respectively. Inversely, protozoal infections (diarrhoeal diseases) were associated with rapid ART-initiation (aHR=1.06). The findings on presenting clinical features and ODs aetiological agents are seemingly unique to the present study as previous evidence is scanty. Nonetheless, a multicentre study from Europe examining TTAT-initiation reported that Tuberculosis (TB), a mycobacterial condition, explained a 38% negative association with rapid ART-initiation (i.e., had longer time to ART-initiation) (aHR=0.62, 95%CI: 0.48-0.80), p<0.01 [100]. In a different study from China, TB was 42.80-fold highly likely to be associated with late/delayed ART start [104]. These findings, though the effect size (ES) estimates were larger than the present study, supports the present study’s findings on association of aetiological agents responsible for causing ODs; in this case, mycobacterial causes, on TTAT-initiation. Although negative associations were unexpected, they somehow reflect population’s dependency and reliance on alternative therapies as first interventions to ill health including HIV other than modern medicine; – a widespread practice in most African populations [120–122].

HIV surrogate markers: CD4+ cell count and VL levels, both demonstrated clinically expected findings. CD4 cell count is an HIV surrogate and prognostic marker of disease progression whereas viral-RNA (VL) is a marker of HIV disease activity [123–126]. In this study, low CD4 count of <250 cells/µL, was 15% positively associated with early (rapid) ART-initiation [aHR=1.15]; a finding which compares with, though lower than 31% from US study [105], but higher than studies from Canada [115] and Mozambique [109]. A UK study reported that having higher median baseline CD4+ count was associated with delayed ART initiation [73] whereas contrarywise, low CD4 count displayed a 2.16-fold to 4.71-fold higher probability of rapid ART-initiation in a study from Italy [127] – much higher that the finding in the present study. Comparatively, KwaZulu Natal study reported that having higher CD4+ cell count (of about 200-350 cell/µL) was associated with rapid 9% ART-initiation [102].

The ADROC Study findings also compare favourably with the Icona Study from Italy that described factors associated with rapid or delayed ART-initiation. The Icona Study reported that overall, PLHIV with lower CD4+ count and higher HIV-RNA were highly associated with rapid ART-initiation. Thus, adjusting for different factors; the Icona study demonstrated that compared to having low CD4+ count <200 cell/µL was associated with 4.71-fold higher probability of initiating treatment early [127]. This finding is congruent with the present study’s findings – though with much higher ES. Comparatively, a European study demonstrated that high CD4+ count (≥200 cells/*µ*L) was correlated to longer TTAT-initiation by 60% [aHR=0.30, 95%CI:0.20-0.44] [100].

However, HIV-RNA load showed negative association to rapid-ART-initiation. The findings demonstrated that high, VL>1,000 copies/mL was 17% negatively associated with rapid ART-initiation and acted as barrier (aHR=0.87). This finding, however, partly reflects previous recommendations that guided clinical practice from the introduction of ARVs. Thus, while HIV-seropositivity regardless of VL level, is currently a universal treatment-initiation eligibility criteria under the UTT strategy, earlier guidelines were contrariwise, based on meeting clinical eligibility criteria of AHD [7, 128, 129]. As a marker of response to ART, viral loads were mostly used to monitor therapy effectiveness following initiation [7]. The present UTT strategy of initiating ART early with definitive lag-time references [98], is the update of initial recommendations of commencing ARVs during “early” HIV infection; where “early” was defined as period up to 6 months after HIV diagnosis [130–133]. In line with current global recommendations, Malawi adopted the UTT strategy in 2016 and embedded it in her successive “Integrated clinical guidelines for HIV management” [134, 135]. The present study findings on VL are consistent and similar to previous evidence elsewhere. The US multicentre study reported that overall, VL was associated with higher adjusted relative risk (ARR) of 23% to 89% of delayed ART-initiation across both early– and late– ART periods [105]. Similarly, a Canadian study identified that high viraemia was highly associated with late-ART-initiation (aOR=5.57) in multivariable logistic regression [35]. Furthermore, another Canadian study on influences of socio-demographic and clinical factors on early ART-initiation reported no significant association of VL with early ART-initiation; inversely translating that high VL was negatively associated with and acted as barrier to rapid ART-initiation [115]. Contrastingly, in the Italian Icona Study, PLHIV with VL (HIV-RNA) ≥100,000 c/mL had a 2.31-fold higher probability (95%CI: 1.37-3.92) of rapid ART-initiation compared to those with VL<100,000 c/mL [127].

This study also demonstrated significant positive association between HIV diagnosis year variable and time-to-ART-initiation. Overall, the predictor was associated with rapid ART-initiation with progressively stepwise pattern increasing trends (rates) with every passing calendar-year except 2007/2008 (Fig. 4). These findings highlight and reflect Malawi’s evolving HIV management over the years since 2004’s initial recommendations. Thus, the progressive trends in time-to-ART-initiation in the present study reflect Malawi’s evolving treatment guidelines over time in response to WHO recommendations: i.e., from David Ho’s “hit early, hit hard” model, to the “treat early and treat hard” strategy in early 2000s based on AHD – WHO clinical stages III and IV, evolving to initiating ART due to low CD4 count below acceptable thresholds; thus, from CD4 ≤200, ≤350, ≤350+TasP, ≤500 cells/µL, and to the present “treat-all” HIV+ persons; the UTT strategy – reflecting the 2000, 2006, 2010, 2013, and 2015 guidelines [136–139]. The findings in the present are consistent with observational cohort results in Vancouver (Canada) which showed that recent annual HIV cases were associated with rapid (early) ART-initiation [101] and are equally consistent with San Francisco Citywide Study results [140]. The San Francisco study, like the (present) ADROC Study, demonstrated a progressive increasing trends and probability of early (rapid) ART-initiation with every passing calendar year, thus; increasing aOR year on: 2014 by 3.0-fold, 2015 by 6.37-fold, 2016 by 12.33-fold, 2017 by 16.84-fold [140] – a clear similar pattern observed in present study. Additionally, HIV diagnosis year variable was a significant finding from multicentre study where compared to 2000-2008 year of diagnosis, 2009-2012 had 2.07-fold (95%CI:1.58-2.72, *p<*0.01) higher probability of rapid ART-initiation [100]; a pattern similar to present one. Nonetheless, present study’s findings are different from Canada’s Montreal study which reported negative association between rapid-ART-initiation and new HIV cases [115].

## CONCLUSION

This study has highlighted insights into factors that have been associated with both early (timely) or late (delayed) initiation of antiretroviral therapy among adults living with HIV in Malawi in the wake of free anti-HIV treatment in the era of clinical eligibility criteria guidelines. The study has identified different demographic, geographic, socioeconomic, and (bio)clinical factors that influenced ART-initiation in among the enrolled cohort besides meeting eligibility criteria. These factors have contributed in some way towards mortality and morbidity from HIV/AIDS in Malawi. Early ART start has been linked with double benefits of as-treatment-as-prevention. Hence, late ART start will obviously put the trajectory in reverse. Unfortunately, there are no similar previous studies from Malawi to compare the present findings with – however, factors impacting time-to-ART-initiation have been examined in studies elsewhere and the findings compare variably.

Although the findings in the present study relate to the previous (and outdated) guidelines on HIV management, the identified factors are still relevant even during the present-day practise and cannot be ignored. Even though Malawi adopted WHO’s recommended UTT strategy of HIV management in 2016, there are still considerable delays in presentation to care, HIV diagnosis, and ART-initiation as well as adherence to treatment that will potentially affect the overall long-term survival outcome. Besides, it has already been established that PLHIV may start and stop HIV care or ART multiple times over the course of their lives, as such, a knowledge of factors for or against early treatment initiation cannot be ignored, overlooked, or overemphasised.

### Strengths and limitations

In this cohort, the relatively larger sample size provides the findings that are to a greater extent representative of adults living with HIV in Malawi. In addition, the data used cut across all three regions of Malawi; South, Central, and Northern regions, and covers all three levels of healthcare establishments where diseases including HIV are treated, namely primary level, secondary tier, and tertiary tier health facilities, making the data fit of a nationally representative sample. The nationally representativeness of the data gives the findings credence, validity, and strength to be applicable and generalisable across Malawi and other areas within the Sub-Saharan African region with similar settings. Hence, the results are relevant especially to Malawi. However, the time-to-ART-initiation descriptions used in the study are rather too generic, broad, and too general and are without reference to any lag-time as current literature stipulates. While this might potentially cause a bit of confusion to some, it ought to be noted that this description reflects previous HIV management guidelines used as reference guide when all enrolled patients in this study commenced ART where lag-times were not applied.

## Data Availability

All data generated or analysed during the current study are available from the corresponding author, HHS, on reasonable request.

## Authors’ contributions

HHS contributed 100% of research work, thus; Conceptualization, Data curation, Methodology, Formal analysis, Writing – original draft, Writing – review & editing, Writing – final draft.

## Acknowledgements

The author is grateful to all gate keepers and research teams from all 8 research sites: Nkhoma Hospital, Lighthouse Trust & Martin Preus Centre, Partners-in-Hope, Mzuzu Hospital, DREAM Centre, Chiradzulu Hospital, QECH, and Thyolo Hospital ART facilities for their undivided support towards this study.

## Availability of data and materials

All relevant data generated or analysed during the current study are available from the corresponding author, HHS, on reasonable request.

## Ethical approval and Informed consent to participate

Study’s ethical approval was granted by the University of Warwick Research Ethics Committee; The Biomedical and Scientific Research Ethics Committee (BSREC) [BSREC approval number: REGO-2015-1576], and from Malawi Research Ethics Committee; The National Health Sciences Research Committee (NHSRC) in the Ministry of Health [NHSRC approval number: NHSRC # 16/3/1555]. The need for consent to participate was waived off due to nature and source of data.

## Supplementary materials

None.

